# Practice-level variation in the provision of subsidized dental services to adult Danes in 2019: A register-based study

**DOI:** 10.1101/2025.02.07.25321838

**Authors:** Eero Raittio, Vibeke Baelum

## Abstract

**Objectives:** The aim was to investigate practice-level variation in common dental diagnostic, preventive, and care services provided for Danish adults who underwent a dental examination.

**Methods:** This was a nationwide register-based study. Subsidized dental services delivered during the 13-week-period subsequent to the provision of one of three eligible dental examinations (extended, basic, or recall examination) during the first nine months of 2019 were investigated. Bayesian multilevel regression models were used to estimate the practice-level average predicted probability of supragingival care, subgingival care, individual prevention, bitewing radiographs, and endodontic treatment, and the average predicted count of extractions and direct restorations while adjusting for individual sociodemographics and dental treatments received during the previous 10 years.

**Results:** The final sample included 445,516 examinations conducted in 1,593 dental practices. Supragingival care after basic or recall examinations showed the lowest practice-level variation, with around two-fold difference between top and bottom 2.5%. Individual preventive services after recall examinations showed the highest variation with over 30-fold difference between top and bottom 2.5%. All other outcomes showed around 3- to 8-fold differences between practices at top and bottom 2.5% across all examination types. The differences across practices were smaller—1.2- to 3.0-fold—when the top 25% and bottom 25% were compared instead.

**Conclusions:** This study found considerable variation in diagnostic, preventive, and treatment services provided for Danish adults who underwent a dental examination. The findings highlight the need for research that can inform evidence-based practice through the development of quality clinical practice guidelines, continuing education programs, and closer surveillance of care delivery.

## Introduction

In dentistry, as well as in medicine, there is considerable evidence of variations in clinical diagnoses and treatment decisions when identical/similar clinical cases are assessed,^1–6^ along with variation in care across countries^7,8^ and regions within countries,^9,10^ and among healthcare providers in real-world settings.^11,12^ Timely diagnosis, prevention, and treatment in dentistry are regarded as crucial. However, each method for diagnosis, prevention, or treatment carry potential benefits and harms, necessitating their thoughtful and judicious application under considerable uncertainty that may stem from limited or poor quality evidence,^13^ or from diagnostic uncertainty.^12^ Both limitations leave room for a considerable provider impact on the care decisions,^12^ and provider characteristics such as experience, skills, diligence, tolerance for uncertainty, knowledge, beliefs about treatment utilities, treatment preferences, diagnostic techniques used, outlier experiences, practice busyness, and adherence to guidelines, are some of the factors that have been proposed as influential.^14–16^ While surprisingly little is known about their relative contribution to variation in service provision, the available evidence points to a considerable contribution of provider-related drivers of the variation in the care provided.^15,17^

These variations represent one pathway through which under- or overdiagnosis, and consequently under- or overtreatment of dental caries and periodontal conditions can occur, issues that have been subjects of long-standing discussions worldwide.^18–23^ Variations among clinical dental practices are also regularly caught and discussed in lay media^24,25^ and have received attention from policymakers and public authorities aiming to maintain quality and equality of the health care system.^26^ It is thus clear that if “two dentists consistently provide a different set of preventive and treatment procedures for patients with similar conditions, then one dentist must be providing less effective care than the other, unless the care leads to equivalent results for patients when compared across a wide range of possible outcomes.”^1^ However, even if the clinical outcomes may not differ, a different set of preventive and treatment procedures are likely associated with different burdens to patients, including costs, time commitments, physical and psychological discomfort. These differences also create different burdens for society, particularly when services are subsidized.

Since 2015, dentists working in Danish oral health care system for adults have been required to use one of three types of clinical examinations as the basis for an assessment of the optimal recall interval for patients depending on their current disease status and risk of new disease development.^27^ The extended diagnostic examination is a first-time examination given to new patients presenting with substantial disease activity and complex treatment need, or to regular patients with sudden manifest disease activity owing to special general or local health conditions. The basic diagnostic examination is also a first-time examination, given to new patients who do not present with the conditions warranting the extended diagnostic examination. The contents of the extended and basic diagnostic examinations are the same, comprising anamnesis taking, examination, recording observations, diagnosis, treatment planning, risk evaluation, determination of recall interval, and general prevention (information), but the fee for the former is about twice that of the latter. The status examination (recall examination) is given to regular patients participating in the recall system, and entails an update of the previous asssement (anamnesis, examination, record taking, diagnosis, treatment planning, risk evaluation, determination of recall interval, and general prevention). The fee for this recall examination is the same as for the basic diagnostic examination.

The actual treatments carried out following diagnosis and treatment planning may include disease intervening services, as well as rehabilitation services. While the former services (restorative treatments, endodontics, periodontal treatments, tooth extractions, individual prevention) are publicly subsidized, and therefore tallied in the National Health Insurance Register, rehabilitation services (crowns, bridges, implants, dentures, splints) are not subsidized and therefore not recorded in the register. While it is plausible that much of the current variation in the oral health care service provision for adults may relate to the rehabilitation aspects of dentistry, it is nonetheless of great interest to explore the variation in the disease intervening services. To quote a former chair of the British Dental Association: “*If we are ever to conquer out twin scourges – caries and periodontal disease – it will probably be as a result of effective prevention and not as a result of intervention with an air rotor, no matter how clever.*”^28^

It is therefore of both scientific and public health interest to examine the practice-level differences in the use of common diagnostic, preventive, and treatment procedures. Such analyses are fundamental for attempts to drive improvements in quality, e.g., through education and regulation, eventually benefiting both patients and society in Denmark. The Danish regions, which administer the dental care for adults, attempt to control practice-level variation through so-called control-statistics, and practices that deviate from the national average by 25% or more may be required to account for their use of specific services and a maximum may subsequently be set just as a refund may be required by the Danish regions. Here, the findings are presented on the practice-level variation in common dental diagnostic, preventive, and care services provided for Danish adults who underwent a dental examination during the first nine months of 2019.

## Material and Methods

The data used for the present analysis originates from a large Danish register-based dynamic cohort study. People entered the cohort in the year they reached the age of 20 years in the period from January 1, 1990, to December 31, 2021 and were permanent residents of Denmark. Via pseudonymized individual identification numbers assigned to all residents of Denmark in the Civil Registration System, information the Educational Register, the Income Statistics Register, the National Health Insurance Service Register and the Register for Selected Chronic Diseases were linked. Dental practices were identified using the health care provided identification code issued by the Danish Regions responsible for organizing health care and dental care for the citizens.

Register-based studies do not require individual informed consent in Denmark, and the study was approved by the Danish Data Protection Agency.

### Outcome window

The National Health Insurance Service Register is an administrative register containing information on all patients, providers and all subsidized treatment codes in Denmark. First, utilizing 2019 data, all patients in dentist practices (specialization code 50) were identified who had received either 1) an extended diagnostic examination (code 1111), indicated for new patients with high disease activity or complex treatment needs, or if high disease activity or complex treatment needs is caused by sudden manifestation of general disease; 2) a basic diagnostic examination (codes 1112 and 1113) indicated for new patients without high disease activity or complex treatment needs; 3) a status examination (codes 1114 and 1115) indicated for updating the status and planning of necessary preventive and therapeutic interventions.

The register lacks specific dates for the treatment delivery but contains information about the week when providers invoiced the regions for the delivery of the subsidized treatments. Most of these invoices are registered on the last week of the month. Accordingly, it is reasonable to assume that the services listed in the invoice were delivered during that week or the three preceding weeks of the month. Thus, separately for each of the three types of examinations, the first week of 2019 when the patient received the examination was identified, and then they were followed up for an additional 13 weeks to detect the treatments delivered to the patient by the same provider during this approximately 3-4-month period. This timeframe for the outcome window was selected to allow sufficient time for providers to address all identified treatment needs from the examinations. To maintain equal length of the outcome window after all examinations, only examinations which took place before the end of September 2019 were included (Figure 1).

**Figure 1.**
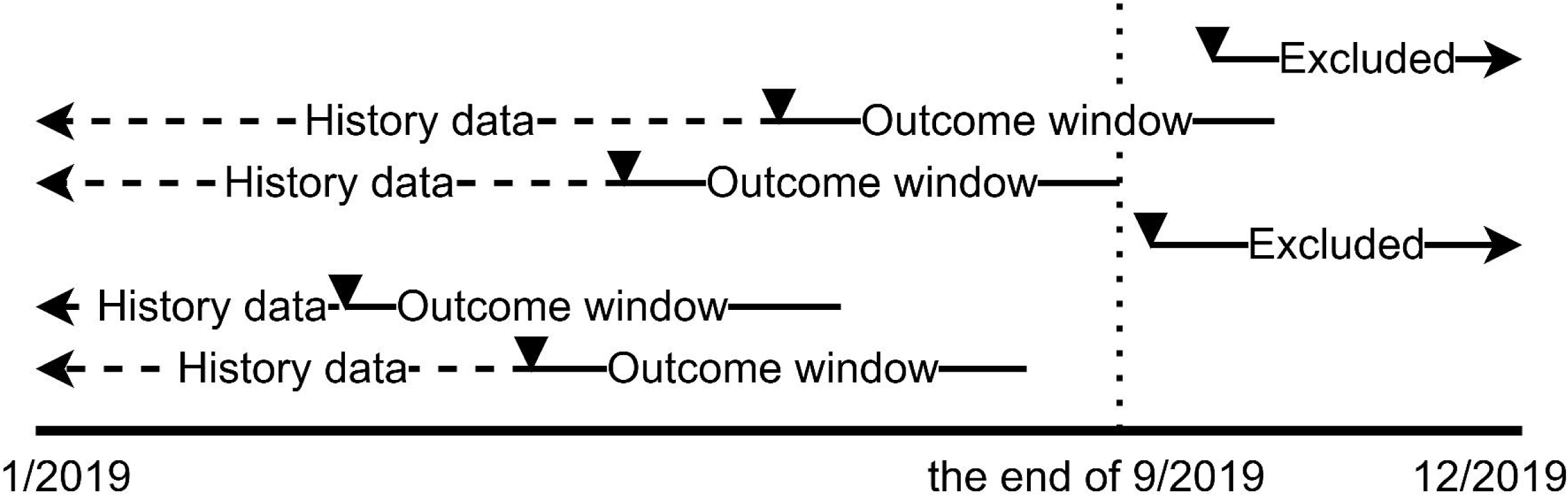
Outcome window. Triangles represent time when one of three eligible dental examinations occurred.

While an individual could thus appear in the dataset for one or more of the three examination types (e.g., when changing provider or in case of sudden manifest disease activity), only the first examination of each type was considered in the analyses.

### Outcomes

Using the data on subsidized dental services delivered during the outcome window period, five binary outcomes reflecting the provision of diagnostic, preventive, and treatment services were formed: 1) supragingival care (including scaling and polishing); 2) subgingival care (excluding periodontal surgery); 3) individual prevention (including oral hygiene advice, fluoride application, smoking, or dietary advice); 4) bitewing radiographs; 5) endodontic treatment. Additionally, two count outcomes were examined: the number of direct restorations (amalgam or composite) and the number of extractions (surgical and nonsurgical).

### Covariates

To take account of differing case-mix across practices, individual level covariates representing sociodemographic characteristics and the dental treatments received from the beginning of 2009 to the beginning of outcome window in 2019 were used to adjust for these factors. Based on the 2019 information, the following covariates were used: age, gender, origin (Danish/immigrant/descendant), region or municipality of residence (15 levels), the highest educational attainment (9 levels), income percentile, and diabetes type 1 or type 2 status (yes/no) (for further detail, see Supplement). Moreover, utilizing data from 2009 up to the individual outcome window in 2019, the following variables were formed and used in the case-mix adjustments: the number of calendar years with at least one dental visit including 1) dental examination, 2) supragingival care, 3) subgingival care, 4) periodontal surgery, 5) individual preventive service, 6) endodontic treatment, or 7) bitewing radiographs. Similarly, the total number of direct restorations and the number of surgical or nonsurgical extractions performed in the period from 2009 up to the individual outcome window in 2019 were calculated and used for the case-mix adjustment. Additionally, to capture time-varying and accumulating effects of income, a variable representing the sum of the annual income percentiles between 2009 and 2018 was generated (for further detail, see Supplement).

### Sample selection

The three final analytic samples, representing each distinct type of dental examinations, were generated by omitting data from individuals who had gaps or incomplete covariate information due to residence outside of Denmark or being under 20 years of age for some part of the period 2009-2019. Data were also omitted for practices that conducted 10 or fewer of the relevant types of examinations to ensure more stable and reliable practice-level estimates. Moreover, to ensure that statistical analyses remained computationally manageable while preserving their validity and generalizability, random samples of 200 status examinations were taken from practices with more than 200 of such examinations. Figure 2 shows the number of practices and individuals that were omitted from analysis for each of these three reasons.

**Figure 2.**
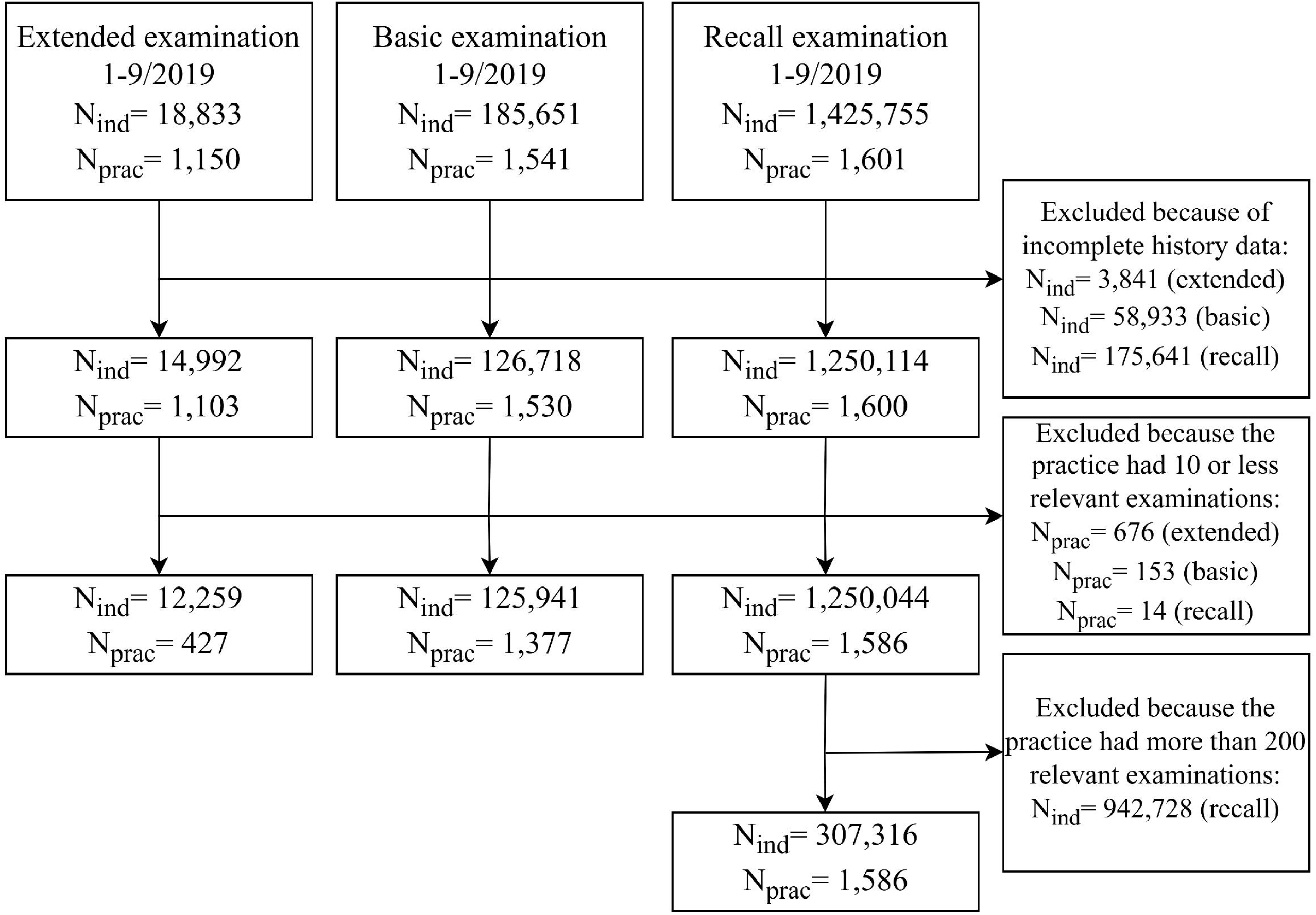
Sample selection process.

### Statistical analyses

Multilevel Bayesian regression models with weak default priors were used to investigate practice-level variation in the outcomes. For binary outcomes analysis was based on logistic regression while count outcomes were analyzed using Poisson regression. First, models were fitted with a random effect for practice but without any individual level covariates to capture the unadjusted practice-level variation in the outcomes. Then, models were fitted which included the random effect for practice and all individual-level covariates as fixed effects. In these models, the variables of age, income percentile, the sum of income percentiles over 2009-2018, and the total number of direct restorations and the number of surgical or nonsurgical extractions were allowed to have nonlinear relationship using natural cubic splines with four knots.

Using the outputs from these regression models, practice-level variation was analyzed by computing the practice-level average predicted outcome probability or count, while keeping individual covariates at their mean or modal values to ensure adjustment for different practice level case-mix. In alignment with data protection rules of the Denmark Statistics, to prevent the identification of individual practices, practice-level predictions were visualized after being grouped into percentiles.

This investigation also involved reporting the median, the quartile values (25% and 75% percentiles) and the top 2.5% and bottom 2.5% of these practice-level average predictions. The ratio between the top 2.5% and bottom 2.5% of these practice-level average predictions was used to express the relative differences between practices with the highest and lowest average predicted outcome probabilities or counts, while excluding the most extreme 2.5% at either end.^29^ Since the Danish regions exert their control statistics based on deviations in excess of 25% from the national average, calculations were also done using the ratio between the quartile values (Table S1).

The analyses were performed in R using mainly tidyverse,^30^ brms^31^ and marginaleffects^32^ packages on Statistics Denmark’s Research Service environment. The study complied with STrengthening the Reporting of OBservational studies in Epidemiology (STROBE) and the Reporting of studies Conducted using Observational Routinely-collected health Data (RECORD) checklists.

## Results

Around 1.5 million adults underwent at least one of the included three types of examinations in around 1,600 dental practices between January and September 2019 (Figure 2). After applying the selection criteria for the sample, the final groups consisted of 12,259 individuals who received an extended examination across 427 practices; 125,941 individuals who underwent a basic examination across 1,377 practices; and 307,316 individuals who attended a recall examination across 1,586 practices (Table 1).

**Table 1.** Characteristics of the final analytic samples for the three types of dental examinations.

|  | **Extended** | **Basic** | **Recall** |
| --- | --- | --- | --- |
| **N (individuals)** | **12,259** | **125,941** | **307,316** |
| **N (practices)** | **427** | **1,377** | **1,586** |
| Age, median (Q1, Q3) | 55 (44, 67) | 51 (40, 64) | 59 (47, 70) |
| Men | 6,194 (51%) | 57,720 (46%) | 140,226 (46%) |
| Origin |  |  |  |
| Danish | 10,794 (88%) | 113,667 (90%) | 291,533 (95%) |
| Immigrant | 1,348 (11%) | 11,003 (8.7%) | 14,345 (4.7%) |
| Descendant | 117 (1.0%) | 1,271 (1.0%) | 1,438 (0.5%) |
| Highest educational attainment |  |  |  |
| Primary school | 3,791 (31%) | 24,153 (19%) | 54,235 (18%) |
| Vocational education | 4,610 (38%) | 44,867 (36%) | 113,977 (37%) |
| Short-cycle higher education | 504 (4.1%) | 6,870 (5.5%) | 17,198 (5.6%) |
| Medium-cycle higher education | 1,619 (13%) | 23,449 (19%) | 60,773 (20%) |
| Long-cycle higher education | 623 (5.1%) | 15,297 (12%) | 36,768 (12%) |
| Other (4 levels) | 1,112 (9.1%) | 11,305 (9.0%) | 24,365 (7.9%) |
| Municipality/region |  |  |  |
| Central Jutland | 1,503 (12%) | 14,585 (12%) | 37,730 (12%) |
| Southern Denmark | 1,690 (14%) | 13,514 (11%) | 33,506 (11%) |
| Copenhagen | 1,093 (8.9%) | 13,882 (11%) | 28,987 (9.4%) |
| Capital region (outside Copenhagen) | 2,622 (21%) | 27,357 (22%) | 76,710 (25%) |
| Zealand | 2,030 (17%) | 19,361 (15%) | 46,188 (15%) |
| Other region or municipality (10 levels) | 3,321 (27%) | 37,242 (30%) | 84,195 (27%) |
| Income percentile, median (Q1, Q3) | 45 (28, 70) | 60 (36, 80) | 61 (36, 83) |
| Diabetes | 1,186 (9.7%) | 6,952 (5.5%) | 17,363 (5.6%) |
| Sum of income percentiles<sup>1</sup>, median (Q1, Q3) | 460 (296, 663) | 556 (361, 747) | 616 (400, 799) |
| Sum of restorations<sup>1</sup>, median (Q1, Q3) | 4 (1, 10) | 4 (1, 8) | 6 (3, 11) |
| Sum of extractions<sup>1</sup>, median (Q1, Q3) | 1 (0, 2) | 0 (0, 1) | 0 (0, 1) |
| Sum of years with endodontic treatment<sup>1</sup>, median (Q1, Q3) | 0 (0, 1) | 0 (0, 1) | 0 (0, 1) |
| Sum of years with individual prevention<sup>1</sup>, median (Q1, Q3) | 1 (0, 2) | 1 (0, 2) | 2 (1, 3) |
| Sum of years with examination<sup>1</sup>, median (Q1, Q3) | 3 (1, 7) | 5 (2, 8) | 9 (7, 10) |
| Sum of years with subgingival care<sup>1</sup>, median (Q1, Q3) | 0 (0, 2) | 0 (0, 1) | 0 (0, 3) |
| Sum of years with periodontal surgery<sup>1</sup>, median (Q1, Q3) | 0 (0, 0) | 0 (0, 0) | 0 (0, 0) |
| Sum of years with radiographs<sup>1</sup>, median (Q1, Q3) | 1 (0, 2) | 2 (1, 3) | 3 (2, 4) |
| Sum of years with supragingival care<sup>1</sup>, median (Q1, Q3) | 1 (0, 5) | 4 (1, 7) | 7 (4, 9) |

<sup>1</sup>= over 2009-2018

Individuals who received recall examinations were, on average, older than individuals who received extended or basic dental examinations (Table 1). Those who received extended examinations were more likely to be immigrants, men, have lower education, lower income, diabetes, and having visited dental care less often and they were likely to have received less examinations and less supragingival care during the previous 10 years than those in the basic or recall examination samples. Differences between individuals in the basic examination and recall examination samples were less clear, but it can be concluded that individuals who were included in the recall examination sample were more likely of Danish origin, older, and having received more restorations, examinations, and supragingival care during the previous 10 years than individuals in the basic examination sample.

Around two-thirds of individuals who underwent an extended or basic examination also had bitewing radiographs taken within the outcome window (approximately 3-4 months), whereas only one-quarter of recall examination patients had bitewing radiographs taken (Table 2). Regarding all three types of examinations, around one half of the individuals received individual prevention. While those who had an extended examination were more likely to receive subgingival care (32%) than those who had basic (20%) or recall (24%) examination, they were significantly less likely to receive supragingival care (33%) than those in the basic (68%) or recall (77%) examination samples. The average number of extractions and restorations received during the outcome window were many fold higher in the extended and basic examinations samples than in the recall examination sample.

**Table 2.** Diagnostic, preventive, and treatments received during the 2019 outcome window by the examination type.

|  |  | **Extended** | **Basic** | **Recall** |
| --- | --- | --- | --- | --- |
| N (patients) |  | 12,259 | 125,941 | 307,316 |
| Bitewing radiograph | N (%) | 7,370 (60%) | 83,498 (66%) | 81,652 (27%) |
| Individual prevention | N (%) | 6,139 (50%) | 63,825 (51%) | 134,263 (44%) |
| Subgingival care | N (%) | 3,956 (32%) | 24,925 (20%) | 73,816 (24%) |
| Supragingival care | N (%) | 4,071 (33%) | 86,007 (68%) | 235,379 (77%) |
| Endodontic treatment | N (%) | 1,630 (13%) | 8,029 (6.4%) | 5,789 (1.9%) |
| Number of extractions | Mean (SD) | 1.26 (3.08) | 0.33 (1.40) | 0.05 (0.34) |
|  | Median (Q1, Q3) | 0 (0,1) | 0 (0,0) | 0 (0,0) |
| Number of restorations | Mean (SD) | 2.06 (3.32) | 1.01 (1.83) | 0.48 (1.04) |
|  | Median (Q1, Q3) | 1 (0,3) | 0 (0,1) | 0 (0,1) |

The multilevel Bayesian regression analyses indicated that, in all three examination types, considerable variability was observed in the average predicted outcome probabilities or counts between practices (Table 3, Figures 3-6). Except for the outcome number of extractions, the adjustment for individual level covariates did not notably reduce the variation across practices. In the following, the adjusted estimates are therefore quoted. The smallest variation across practices (on the relative ratio scale) occurred in the probability of providing supragingival care after the basic or the recall examination (Figure 3B,C). Following these two examination types, practices in the top 2.5% for predicted probability were approximately twice as likely to provide supragingival care compared to practices in the bottom 2.5%. Practices in the top 25% for the predicted probability were approximately 1.20 times more likely to provide this service than were practices in the bottom 25% (Table S1), as also indicated by the flatness of the central portions of the curves in Figure 3B, C. However, this relatively small variation should be seen in the light of the generally high predicted probability of provision of this service, being 70% following a basic examination and 84% following a recall examination (Table 3, Figure 3B,C). In contrast, the ratio between the top 2.5% and bottom 2.5% in average predicted probability of providing individual preventive service after a recall examination was over 30 (Table 3, Figure 4C). There were also over 10-fold relative differences across practices in the probability of providing subgingival care after a recall examination (Table 3, Figure 3F), and in the predicted number of extractions performed after an extended examination (Table 3, Figure 5D). Otherwise, ratios between the top 2.5% and bottom 2.5% of these practice-level average predictions indicated that the top 2.5% had around 3-8 times higher average predicted outcome probability or count than the bottom 2.5% in all examination samples (Figures 3-5 and S1).

**Table 3.**
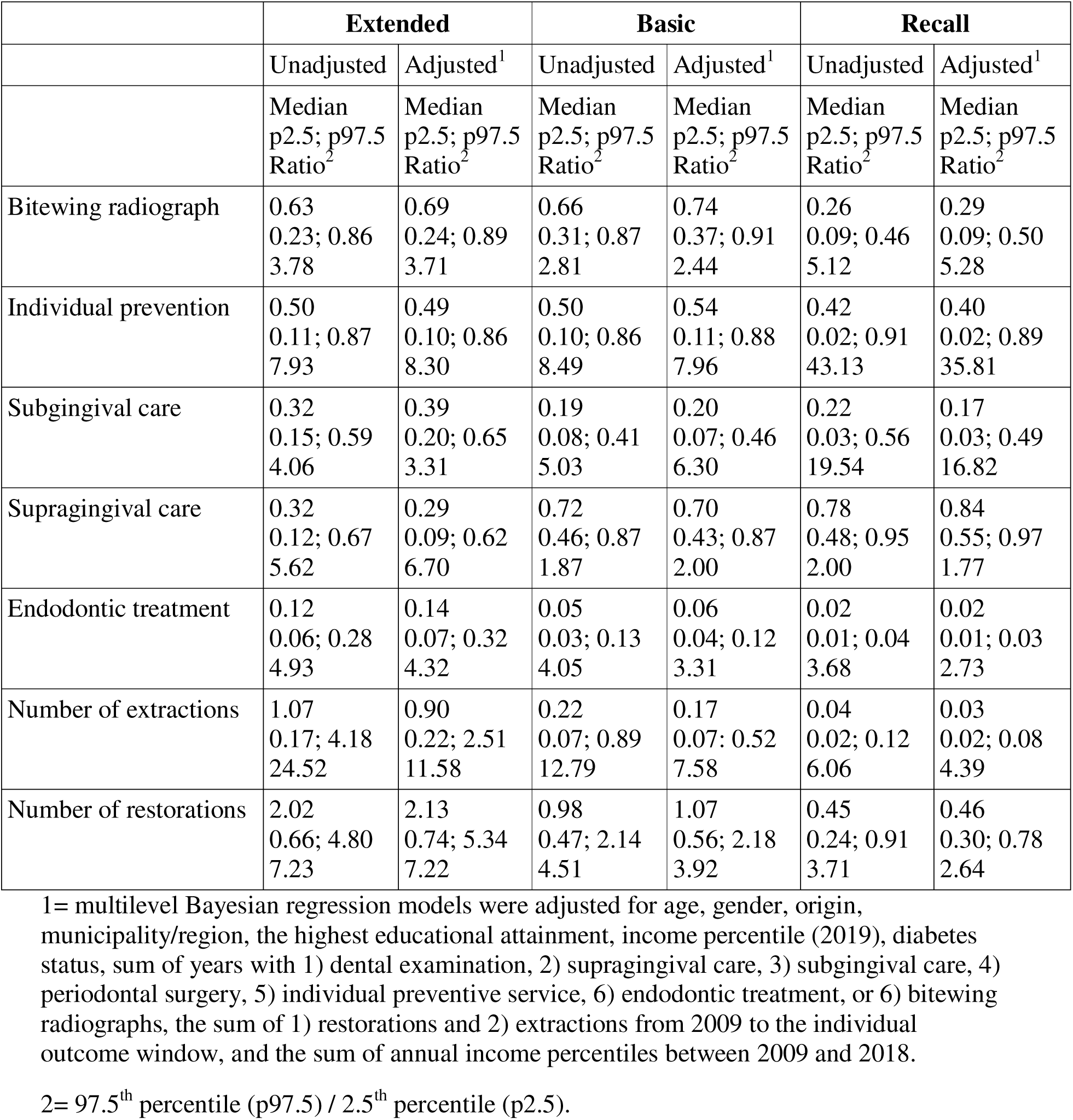
Unadjusted and adjusted average predicted outcome probability or count across practices.

|  | **Extended** |  | **Basic** |  | **Recall** |  |
| --- | --- | --- | --- | --- | --- | --- |
|  | Unadjusted | Adjusted<sup>1</sup> | Unadjusted | Adjusted<sup>1</sup> | Unadjusted | Adjusted<sup>1</sup> |
|  | Median<br>p2.5; p97.5<br>Ratio<sup>2</sup> | Median<br>p2.5; p97.5<br>Ratio<sup>2</sup> | Median<br>p2.5; p97.5<br>Ratio<sup>2</sup> | Median<br>p2.5; p97.5<br>Ratio<sup>2</sup> | Median<br>p2.5; p97.5<br>Ratio<sup>2</sup> | Median<br>p2.5; p97.5<br>Ratio<sup>2</sup> |
| Bitewing radiograph | 0.63<br>0.23; 0.86<br>3.78 | 0.69<br>0.24; 0.89<br>3.71 | 0.66<br>0.31; 0.87<br>2.81 | 0.74<br>0.37; 0.91<br>2.44 | 0.26<br>0.09; 0.46<br>5.12 | 0.29<br>0.09; 0.50<br>5.28 |
| Individual prevention | 0.50<br>0.11; 0.87<br>7.93 | 0.49<br>0.10; 0.86<br>8.30 | 0.50<br>0.10; 0.86<br>8.49 | 0.54<br>0.11; 0.88<br>7.96 | 0.42<br>0.02; 0.91<br>43.13 | 0.40<br>0.02; 0.89<br>35.81 |
| Subgingival care | 0.32<br>0.15; 0.59<br>4.06 | 0.39<br>0.20; 0.65<br>3.31 | 0.19<br>0.08; 0.41<br>5.03 | 0.20<br>0.07; 0.46<br>6.30 | 0.22<br>0.03; 0.56<br>19.54 | 0.17<br>0.03; 0.49<br>16.82 |
| Supragingival care | 0.32<br>0.12; 0.67<br>5.62 | 0.29<br>0.09; 0.62<br>6.70 | 0.72<br>0.46; 0.87<br>1.87 | 0.70<br>0.43; 0.87<br>2.00 | 0.78<br>0.48; 0.95<br>2.00 | 0.84<br>0.55; 0.97<br>1.77 |
| Endodontic treatment | 0.12<br>0.06; 0.28<br>4.93 | 0.14<br>0.07; 0.32<br>4.32 | 0.05<br>0.03; 0.13<br>4.05 | 0.06<br>0.04; 0.12<br>3.31 | 0.02<br>0.01; 0.04<br>3.68 | 0.02<br>0.01; 0.03<br>2.73 |
| Number of extractions | 1.07<br>0.17; 4.18<br>24.52 | 0.90<br>0.22; 2.51<br>11.58 | 0.22<br>0.07; 0.89<br>12.79 | 0.17<br>0.07; 0.52<br>7.58 | 0.04<br>0.02; 0.12<br>6.06 | 0.03<br>0.02; 0.08<br>4.39 |
| Number of restorations | 2.02<br>0.66; 4.80<br>7.23 | 2.13<br>0.74; 5.34<br>7.22 | 0.98<br>0.47; 2.14<br>4.51 | 1.07<br>0.56; 2.18<br>3.92 | 0.45<br>0.24; 0.91<br>3.71 | 0.46<br>0.30; 0.78<br>2.64 |

1= multilevel Bayesian regression models were adjusted for age, gender, origin, municipality/region, the highest educational attainment, income percentile (2019), diabetes status, sum of years with 1) dental examination, 2) supragingival care, 3) subgingival care, 4) periodontal surgery, 5) individual preventive service, 6) endodontic treatment, or 6) bitewing radiographs, the sum of 1) restorations and 2) extractions from 2009 to the individual outcome window, and the sum of annual income percentiles between 2009 and 2018.
2= 97.5<sup>th</sup> percentile (p97.5) / 2.5<sup>th</sup> percentile (p2.5).

**Figure 3.**
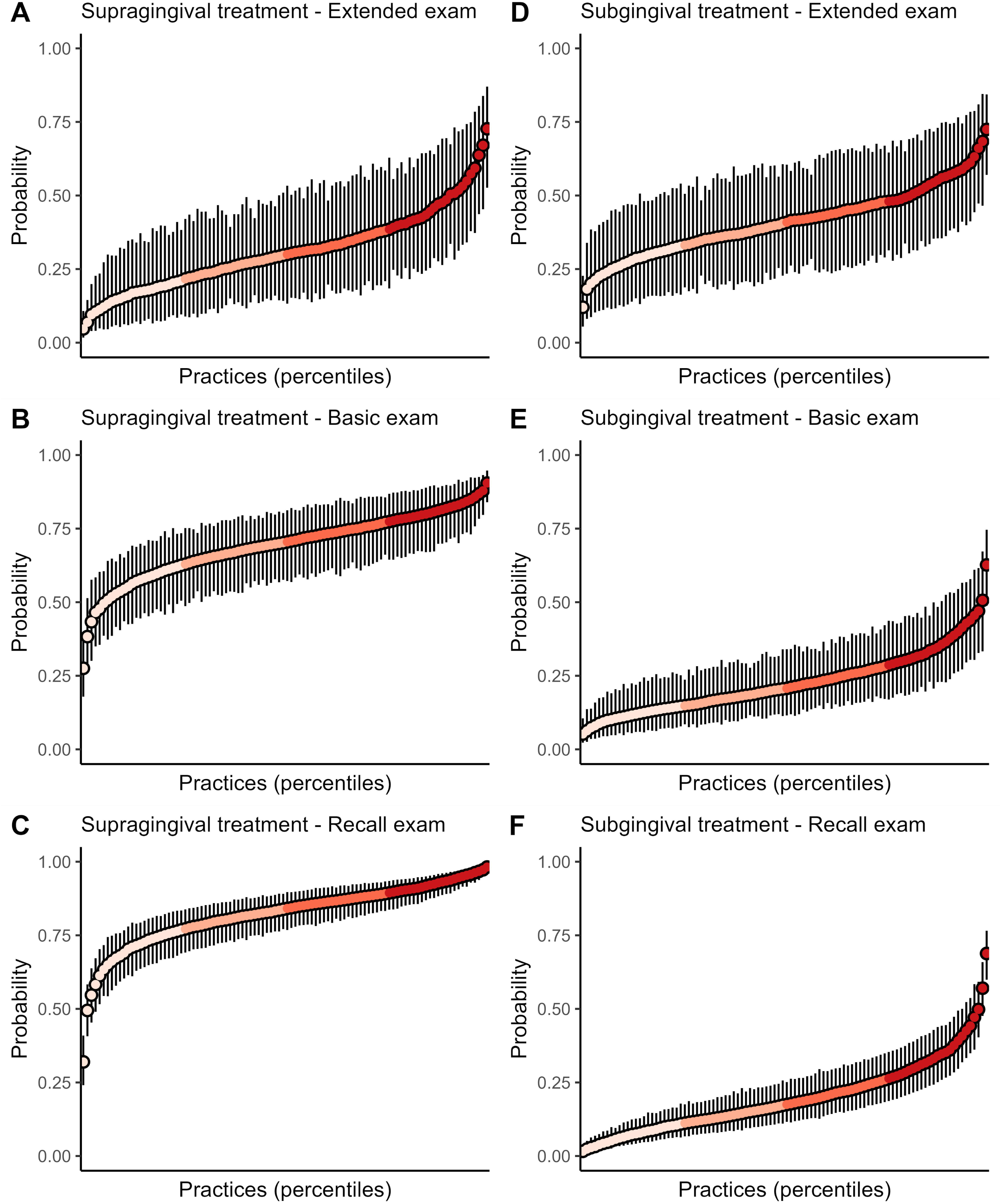
Average predicted probability of supragingival (A-C) and subgingival treatment (D-F) with 95% credibility intervals across practice percentiles for each type of examination. Predictions were generated using multilevel Bayesian regression models, with individual-level covariates held at their mean or modal values.

**Figure 4.**
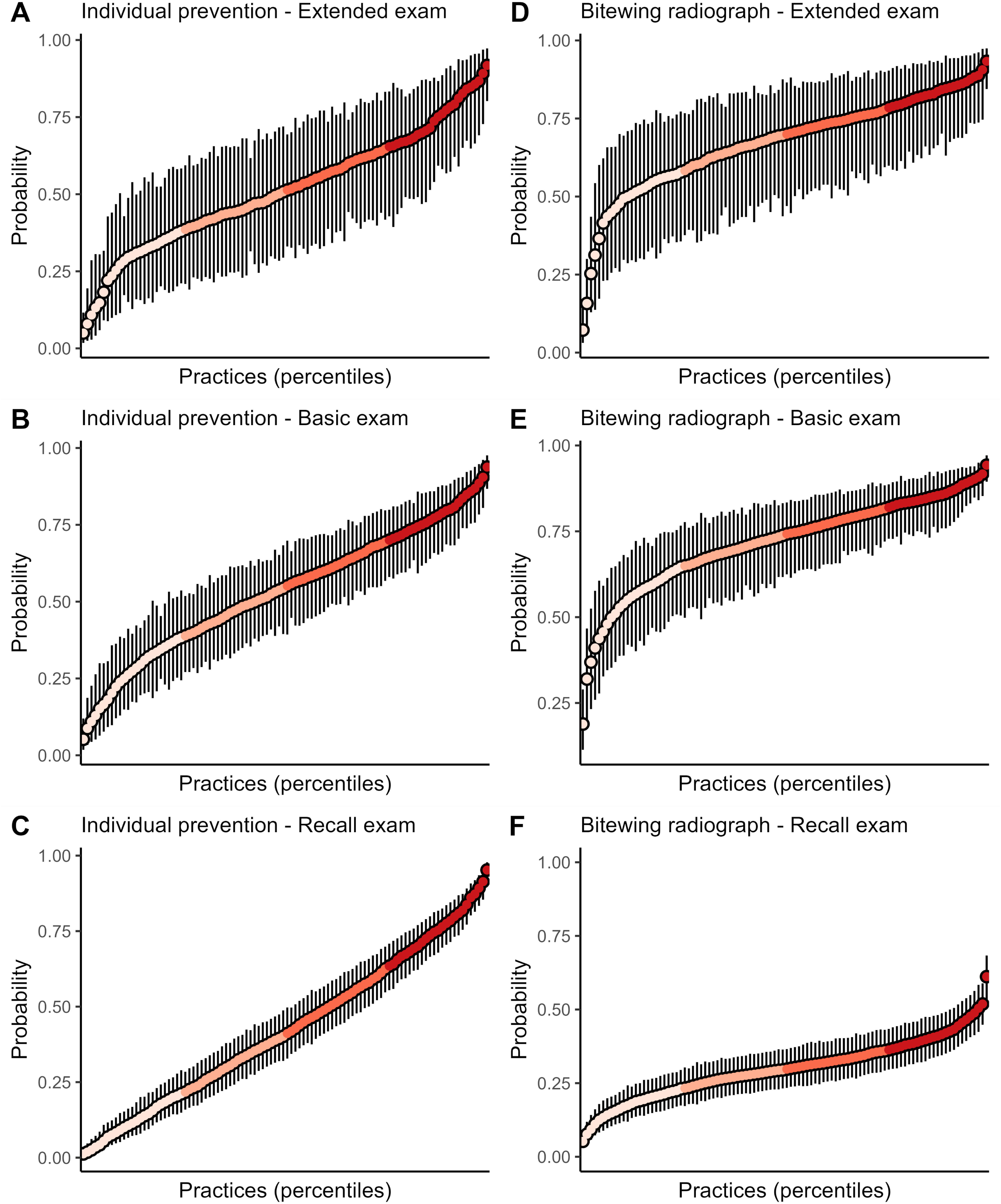
Average predicted probability of individual prevention (A-C) and bitewing radiographs (D-F) with 95% credibility intervals across practice percentiles for each type of examination. Predictions were generated using multilevel Bayesian regression models, with individual-level covariates held at their mean or modal values.

**Figure 5.**
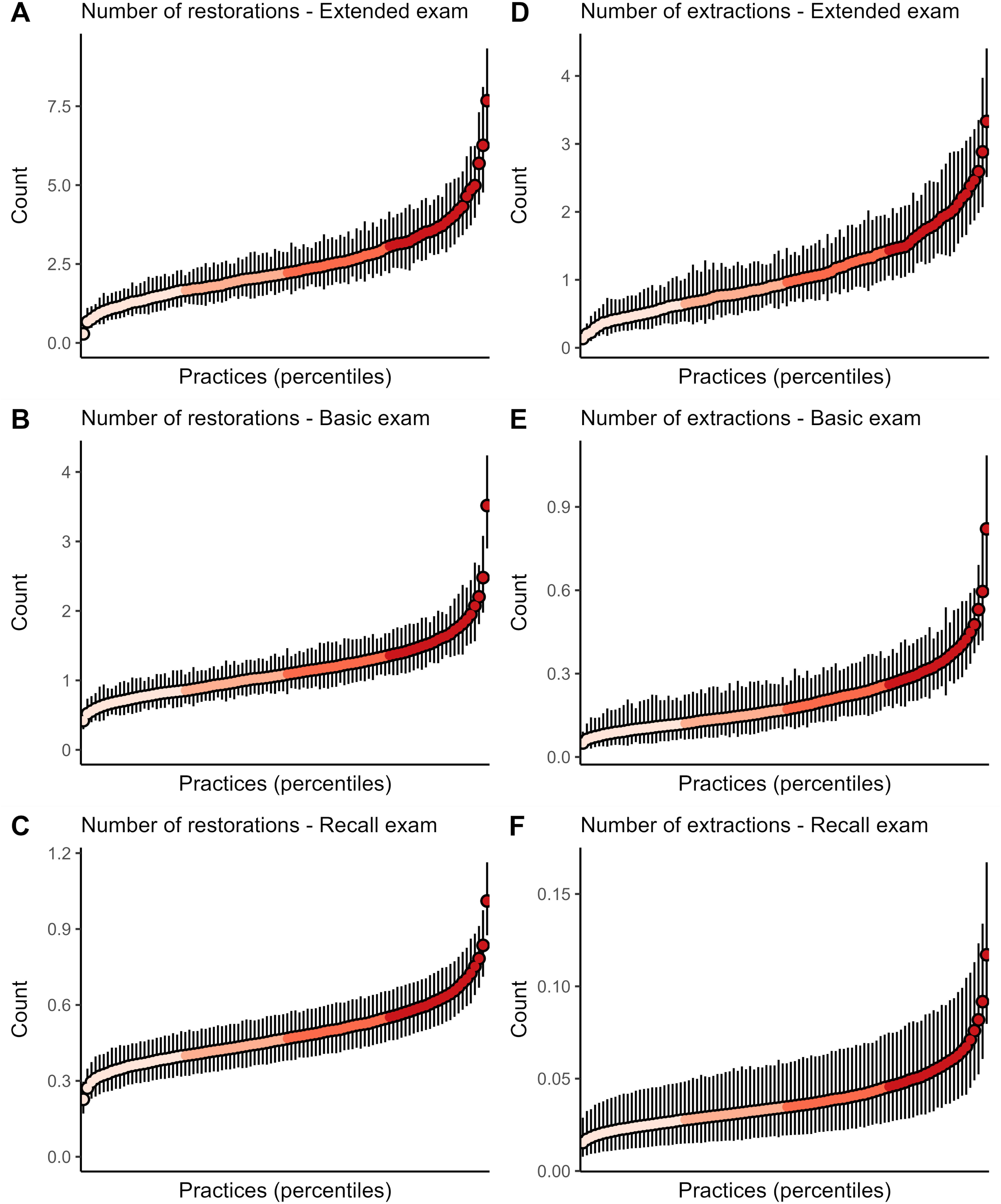
Average predicted count of restorations (A-C) and extractions (D-F) with 95% credibility intervals across practice percentiles for each type of examination. Predictions were generated using multilevel Bayesian regression models, with individual-level covariates held at their mean or modal values.

## Discussion

This study investigated practice-level variation in dental services for Danish adults who underwent one of three different dental examination types. Substantial variability was observed across practices in the provision of diagnostic, preventive, and restorative services. Comprehensive adjustment for individual-level covariates (including demographic and socioeconomic factors, as well as 10-year history of dental treatments) did not considerably explain the variation across practices. The variation across practices occurred across all examination types. Overall, the provision of the investigated diagnostic, preventive, and treatment services was around 2- to 8-fold higher in practices in the top 2.5% than in the bottom 2.5%. The highest variation across practices occurred in providing individual prevention (36-fold) and subgingival care (16-fold) after a recall examination, and in the number of extractions (12-fold) following the extended diagnostic examination for patients with high disease activity or complex treatment needs. When comparing the top 25% and bottom 25% instead, the differences across practices were smaller, ranging from 1.2- to 3.0-fold.

Clearly, some of the variation in service provision could be explained by some practices offering services as referral practices for e.g., periodontal therapy or endodontics. In Denmark, only two specializations exist (orthodontics and oral and maxillofacial surgery), and the referral practices are therefore clinics in which one or more of the associated providers have acquired alternative competences within the area of referral. Such practices will necessarily be characterized by higher levels of service provision within their respective expertises. These practices are not officially registered and it is therefore not possible to exempt them from the analysis. However, as they are few, they are unlikely to substantially affect the analyses. Hence, as of late 2024, 22 Danish practices offered referral services for endodontics, while 11 offered referral services for periodontal therapy.^33^

The observed practice-level variation must necessarily be seen in the light of the level of service provision. It is thus noteworthy that whereas sub- and supragingival care was almost equally likely after an extended examination, as indicated by the average predicted probabilities of 39% and 29% (Table 3, Figure 3A,D), this contrast was reversed after a basic examination (70% and 20%, Figure 3B,E) and even accentuated after the recall examination (84% vs 18%, Figure 3C,F). Considering that service provision was generally lower after a recall examination than after the other two examination types, this indicates that the provision of supragingival treatment to 84% of the patients after a recall examination might represent a sales-compensating act. The relatively limited variation observed in the provision of supragingival therapy following both the recall examination and the basic examination further indicates that the dental practices generally agree on a practice involving a high frequency of provision of this service, even though its effectiveness in preventing tooth loss is quite limited even when delivered on annual basis or or more frequently in Denmark.^34^

The frequency of provision of bitewing radiographs following a recall examination was surprisingly high, with radiographs being provided to a median of 29% percent of the recall patients. Combined with the highly variable frequency of provision of the individual prevention service, this may indicate that treatment is often focused on the detection of carious cavities, which may be seen in bitewings when sufficiently deep, rather than on the control of early caries lesions, which do not show on radiographs^35–37^ but can be managed non-operatively.^38,39^

In view of the indications for the different types of examinations, it was remarkable that the level of provision of individual prevention services was quite uniform across examination types. In particular, one might have expected a substantially higher level of provision of this service following the extended examination which is indicated for new patients presenting with substantial disease activity and complex treatment need, or regular patients with sudden manifest disease activity owing to special general or local health conditions. It is also surprising that 44% of patients had received individual preventive services after recall examinations, even though the majority of these patients had been examined annually, or almost annually, during the last 10 years (median: 9 years, Q1: 7 years, Q3: 10 years; Table 1). These patients only rarely needed extractions or endodontic treatments after the recall examination. According to legal regulations, individual preventive services should be provided for patients with active caries, gingivitis, periodontitis and peri-implantitis, or other dental disease requiring preventive treatment. These findings may thus raise questions about the effectiveness of these regular recall examination visits in reducing the need for individual preventive services. In addition, because the meaning of active oral disease is not defined in the regulation, the findings may highlight the interpretational slack relating to the perceived need for the individual preventive service. Hence, for instance, if any sign of gingival inflammation, is taken to indicate a need for the individual preventive service, e.g. to prevent periodontal attachment loss,^40^ it is clearly possible to provide this service to essentially all recall patients, as epidemiological findings suggest that bleeding on probing is almost ubiquitous in the adult Danish population.^41^ Finally, the high level of provision of individual preventive services to recall patients might reflect interpretational variations regarding the precise activities needed to give grounds for charging for this service code. Therefore, the contents of the individual preventive service entails demonstration of the signs of disease and of plaque deposits, individually tailored oral hygiene instruction, fluoride treatment of active caries lesions and professional plaque control (polishing), and it is clearly possible to exert some flexibility in the interpretation of these activities.

The Danish regions, under whose jurisdiction the Danish dental services for adults fall, have tried to control the variation between providers in the service provision through the use of so-called control-statistics. Hence, if a provider turns out to either exceed or fall behind by 25% or more from the national average of the number of services provided per patient, taking into account the age and gender distributions of the patients, the region may explore the reasons for such deviations.^42^ These reasons can, as mentioned earlier, be legitimate (e.g., in case of a referral practice), but they may also be found to be signs of problematic standards, in which case the provider may be issued with a warning or have a maximum number of services imposed. This control system has been in place for many years, and until the end of 2015 possibly problematic practice standards were defined as deviations of 40% or more. However, from 2016 these limits were narrowed to deviations of 25% or more. The results of the present study, which has employed a much more elaborate adjustment for the patient case-mix differences between the practices compared, indicate that there is still room for improvements with respect to the variation in service provision.

It was not possible to account for patient preferences and values possibly influencing the service provision. However, patient values and preferences would be an important and acceptable, not to say preferable, source of variation across practices. Previous studies have shown that dentists tend to be somewhat hesitant to implement shared decision-making in a clinical context,^43–46^ and it is not known to what extent care provision is really shaped by patient preferences and values. Moreover, it is likely that patient views are weighed differently for diagnostic, preventive, or more invasive procedures (like extractions), making it challenging to speculate how high the variation across practice would have been had it been possible to adjust for patient preferences and values. Presumably, there would still have been considerable variation in clinical practice across providers. Similarly, because studies on identical cases^1–6^ have shown that there exists considerable variation in diagnostic and treatment practices among professionals, it is likely that there would still have been considerable variation across practices, even if it had been possible to adjust more accurately for patient dental and general health status. Accordingly, the comprehensive adjustment set employed here did not considerably explain the variation across practices.

Other limitations of the study that deserve mention include some uncertainties regarding the timing of services. Due to not having the actual dates of treatment delivery in the register, there may be some uncertainty about the temporal sequence of examinations and services provided within the month of examination. Hence, reimbursement claims are usually sent to the Danish regions at the end of each month and it is therefore possible, but unlikely on a large scale, that some of the services have actually been provided before the examination. In any case, they are still conducted by the same practice. The register data also lacks individual practitioner level information, since clinics/practices are identified by a provider number that covers all dentists working in that practice. This number may vary from a single dentist to several professionals (dentists and dental hygienists), all working under one and the same provider number. It was therefore not possible to identify the individual professionals or their profession, which may also explain some of the variation observed, e.g., in the recall examinations which can be conducted by either dentists or dental hygienists depending on the practice. Considering the bigger dental practices, it is likely that considerable within practice variation may exist, which could thus not be captured or controlled here. Even though the generalizability of these findings to other countries may be limited (e.g. due to different treatment codes and health care systems), it is highly probable that quite similar variations in dental practices occur globally. Therefore, it is essential that patients, professionals, and regulators are kept informed about the extent and scale of these variations at the local level, as well as their trends over time.

The current study found considerable variation in diagnostic, preventive, and care services provided for Danish adults who underwent a dental examination. The findings highlight the need for research that can inform evidence-based practice through the development of quality clinical practice guidelines, continuing education programs, and closer surveillance of care delivery.

## Acknowledgements

This work was partly supported by the Danish Rheumatism Association (Gigtforeningen) #R171-A5894.

## Author contributions

Eero Raittio: Conceptualization; Formal Analysis; Methodology; Investigation; Visualization; Writing – original draft; Writing – review & editing.

Vibeke Baelum: Conceptualization; Data curation; Investigation; Project administration; Supervision; Writing – original draft; Writing – review & editing.

## Data availability statement

The data that support the findings of this study are available from Statistics Denmark. Restrictions apply to the availability of these data, which were used under license for this study.

## Funding statement

We thank the Danish Rheumatism Association (Gigtforeningen, #R171-A5894) for the financial support.

## Conflict of interest disclosure

The authors declare that they have no known competing financial interests or personal relationships that could have appeared to influence the work reported in this paper.

## Ethics approval statement

Register-based studies do not require individual informed consent. The study was approved by the Danish Data Protection Agency (2015-57-0002) and Aarhus University (2016-051-000001-914).

## Patient consent statement, Permission to reproduce material from other sources and Clinical trial registration

Not required for this type of study.

## Notes

### Competing Interest Statement

The authors have declared no competing interest.

