## Supplementary material for "Practice-level variation in the provision of subsidized dental services to adult Danes in 2019: A register-based study"

**Supplementary information**

**Variables**

**Gender**: Men; Women. Information from: The Danish Civil Registration System (Pedersen 2011). Variable name in the register: KOEN.

**Origin**: Categorized into two groups: 1) Immigrants or descendants, and 2) persons of Danish origin. (Pedersen 2011). Variable name in the register: IE_TYPE

**Region/Municipality of residence**: Municipality if a person lived in one of ten biggest municipalities (Copenhagen, Aarhus, Odense, Aalborg, Vejle, Kolding, Esbjerg, Viborg, Randers, Frederiksberg) and one of the regions otherwise. Information from: The Danish Civil Registration System (Pedersen 2011). Variable names in the register: KOM and REG.

**Highest completed education**: categorized into eight groups:

- Primary school
- High school
- Vocational
- Short-cycle higher education or qualifying exam
- Medium-cycle higher education
- Bachelor
- Long cycle higher education
- Researcher education
- Unknown

Information from the Danish Educational Register (Jensen and Rasmussen 2011, Pallesen et al 2010). Categorized based on the variable in the register AUDD. Qualifying exam and Researcher education categories were combined with closest neighboring category due to their small size.

**Income percentile**: People were divided into 100 groups (percentiles) for each year from 2009 to 2019 based on their total annual personal income. Personal income in total is equal to the sum of business income, transfer income, property income (excluding calculated rental value of own home) and other non-classifiable income that can be attributed directly to the individual. The amount is before tax deduction, labor market contribution and special pension contribution, and interest expenses are not deducted. Information from the Danish Income Statistics Register (Baadsgaard and Quitzau 2011). Variable name in the register: PERINDKIALT_13.

**Dental service variables**

From the National Health Insurance Service Register (Andersen et al. 2011), we used the dental treatment codes that have been used over time since 2009 in the National Health Insurance scheme covering the subsidized dental care for all adult permanent residents in Denmark. According to the descriptions of the codes, services were categorized as follows:

| **Service** | **Codes** | **Details** |
| --- | --- | --- |
| Supragingival treatment | 1301, 1302 | Supragingival periodontal treatment (scale and polish) |
| Subgingival treatment | 1420, 1425, 1430, 1452, 1453, 1431 | Subgingival periodontal treatment (instrumentation) and control |
| Periodontal surgery | 1440, 1454 | Periodontal surgery and control |
| Dental restorations | 1501, 1502, 1503, 1504, 1505, 1506, 1507, 1509, 1551, 1552, 1553, 1554, 1555, 1556, 1557, 1558, 1559 | Amalgam and composite restorations |
| Oral examination | 1111, 1112, 1113, 1114, 1115, 1116, 1140, 1141, 1160, 1170, 1171, 1180, 2910, 1415 | Focused (e.g., periodontal) or general examinations and dental check-ups, includes also general oral hygiene advice |
| Individual prevention | 2920, 2930 | Individualized oral hygiene advice, fluoride application, smoking advice, or dietary advice |
| Bitewing radiograph | 1150, 1151, 1152, 1153 | - |
| Tooth extractions | 1701, 1705, 1801 | Non-surgical extractions and surgical extractions (i.e. with gingival or mucosal incision, root sectioning or removal of bone tissue) |
| Endodontic treatment | 1600, 1601, 1605, 1606 | Root canal treatments, periapical surgery, and pulpotomies |

**Incident diabetes mellitus type 1 or 2:** Data on incident diabetes between 1997 and 2019 comes from the Registry for Selected Chronic Diseases and Severe Mental Disorders (The Danish Health Data Authority 2024). It combines information from the Danish National Prescription Registry (Pottegård A, et al. 2017), including all prescriptions in Denmark, and the Danish National Patient Registry (Schmidt M, et al. 2015) which contains information on all visits, procedures, and admissions to all Danish somatic hospitals, emergency departments, and hospital-associated outpatient clinics.

Individuals were classified as having incident diabetes mellitus (type 1 or 2) in the beginning of 2019 if the person meet one of the following criteria between 1997 and 2018.

Diabetes type 1:

- Individuals registered with at least two purchases of insulin or insulin analogs (A10A, except combination medicines including GLP1-analogues and insulins, A10AE54 or A10AE56) in the Danish National Prescription Registry.
- Individuals registered with a relevant primary or secondary diagnosis (E10, diabetes type I, or its sub-codes under ICD-10) in the Danish National Patient Registry.

Diabetes type 2:

- Individuals registered with at least two purchases of medication aimed at lowering blood glucose (A10B, except A10BJ, A10BK01, A10BK03) or combination medicines including GLP1-analogues and insulins (A10AE54 or A10AE56) in the Danish National Prescription Registry.
- Individuals registered with a relevant primary or secondary diagnosis (E11, diabetes type 2, or its sub-codes under ICD-10) in the Danish National Patient Registry.

Excluded were:
Women who have been exclusively treated with metformin (ATC code A10BA02) and there were signs that they could have polycystic ovary syndrome (prescription for G03GB02, G03HB or diagnosis code E282).

Women who have a code for gestational diabetes (ICD-10 code O24.4) and who have only registered purchase of antidiabetics (A10) within 280 days before first contact or 280 days after last contact with gestational diabetes according to the Danish National Patient Registry

Table S1. Unadjusted and adjusted average predicted outcome probability or count across practices.

|  | **Extended** | | **Basic** | | **Recall** | |
| --- | --- | --- | --- | --- | --- | --- |
|  | Unadjusted | Adjusted^1^ | Unadjusted | Adjusted^1^ | Unadjusted | Adjusted^1^ |
|  | Median  Q1; Q3  Ratio^2^ | Median  Q1; Q3  Ratio^2^ | Median  Q1; Q3  Ratio^2^ | Median  Q1; Q3  Ratio^2^ | Median  Q1; Q3  Ratio^2^ | Median  Q1; Q3  Ratio^2^ |
| Bitewing radiograph | 0.63  0.52; 0.72  1.40 | 0.69  0.56; 0.77  1.37 | 0.66  0.56; 0.75  1.34 | 0.74  0.65; 0.81  1.26 | 0.26  0.20; 0.32  1.57 | 0.29  0.23; 0.36  1.55 |
| Individual prevention | 0.50  0.37; 0.65  1.75 | 0.49  0.36; 0.64  1.78 | 0.50  0.35; 0.66  1.91 | 0.54  0.38; 0.69  1.80 | 0.42  0.22; 0.65  3.00 | 0.40  0.21; 0.62  2.92 |
| Subgingival care | 0.32  0.24; 0.40  1.62 | 0.39  0.32; 0.48  1.51 | 0.19  0.14; 0.25  1.81 | 0.20  0.15; 0.28  1.90 | 0.22  0.15; 0.31  2.09 | 0.17  0.11; 0.26  2.35 |
| Supragingival care | 0.32  0.22; 0.42  1.90 | 0.29  0.20; 0.38  1.91 | 0.72  0.64; 0.78  1.22 | 0.70  0.63; 0.77  1.23 | 0.78  0.70; 0.85  1.21 | 0.84  0.77; 0.89  1.16 |
| Endodontic treatment | 0.12  0.09; 0.16  1.69 | 0.14  0.11; 0.18  1.60 | 0.05  0.04; 0.07  1.60 | 0.06  0.05; 0.07  1.49 | 0.02  0.01; 0.02  1.54 | 0.02  0.01; 0.02  1.46 |
| Number of extractions | 1.07  0.60; 1.81  3.03 | 0.90  0.60; 1.39  2.31 | 0.22  0.13; 0.37  2.77 | 0.17  0.12: 0.25  2.15 | 0.04  0.03; 0.06  1.82 | 0.03  0.03; 0.04  1.63 |
| Number of restorations | 2.02  1.38; 2.75  1.99 | 2.13  1.55; 2.89  1.87 | 0.98  0.76; 1.23  1.63 | 1.07  0.85; 1.33  1.57 | 0.45  0.37; 0.56  1.54 | 0.45  0.40; 0.55  1.37 |

1= multilevel Bayesian regression models were adjusted for age, gender, origin, municipality/region, the highest educational attainment, income percentile (2019), diabetes status, sum of years with 1) dental examination, 2) supragingival care, 3) subgingival care, 4) periodontal surgery, 5) individual preventive service, 6) endodontic treatment, or 6) bitewing radiographs, the sum of 1) restorations and 2) extractions from 2009 to the individual outcome window, and the sum of annual income percentiles between 2009 and 2018.

2= 3^rd^ quartile (Q3) divided by 1^st^ quartile (Q1)

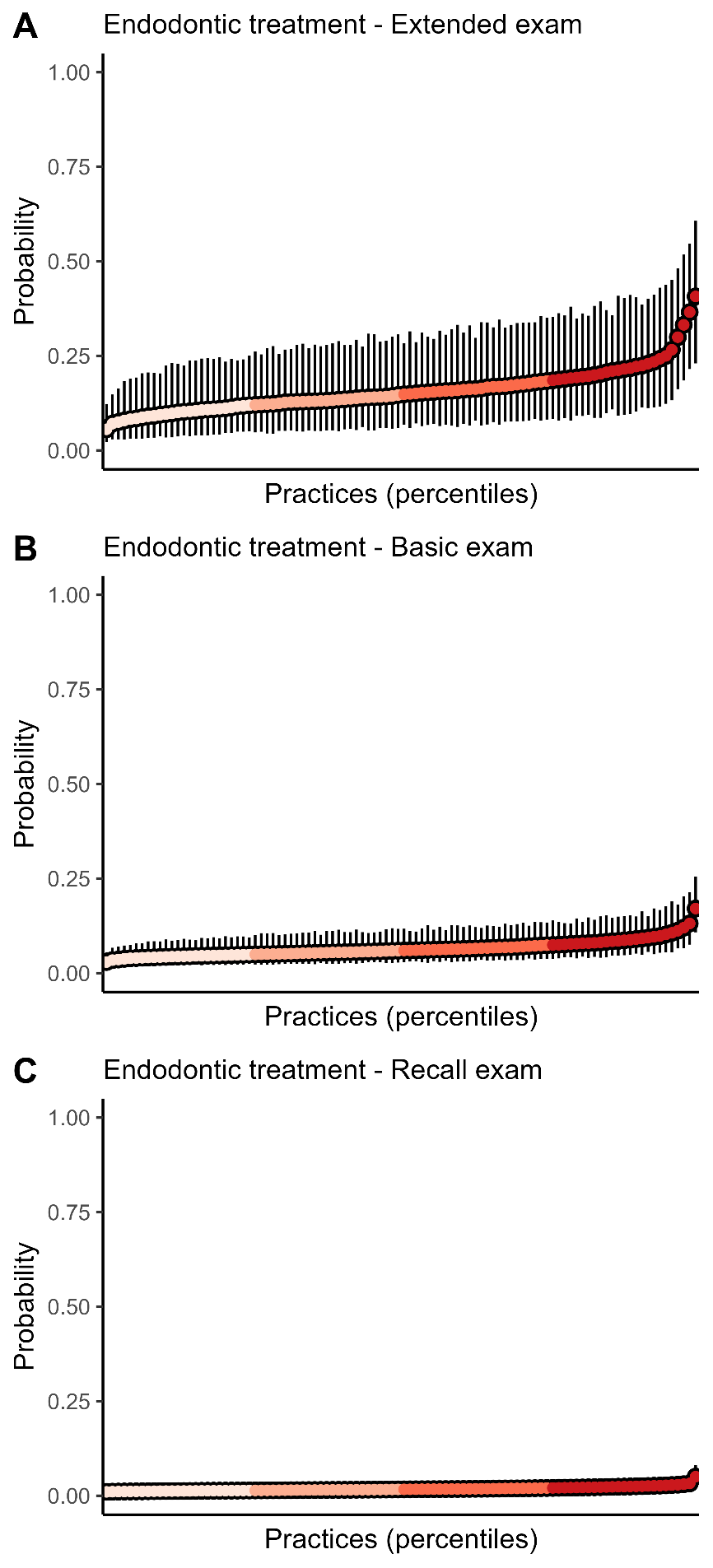

Figure S1. Average predicted probability of endodontic treatment with 95% credibility intervals across practice percentiles for each type of examination (A–C). Predictions were generated using multilevel Bayesian regression models, with individual-level covariates held at their mean or modal values.
